# Modeling SARS-CoV-2 transmission at a winter destination resort region with high outside visitation

**DOI:** 10.1101/2021.08.18.21262227

**Authors:** E.J. Wu, J. Bayham, E. Carlton, J. Samet, A.G. Buchwald

## Abstract

Travel destinations, particularly large resorts in otherwise small communities, risk infectious disease outbreaks from an influx of visitors who may import infections during peak seasons. The COVID-19 pandemic highlighted this risk in the context of global travel and has raised questions about appropriate interventions to curb the potential spread of infectious disease at tourist destinations. In Colorado, the initial outbreaks of SARS-CoV-2 in the state occurred in ski communities, leading to large economic losses from closures and visitor restrictions. In this study, we modeled SARS-CoV-2 transmission during the 2020-21 season in a ski region of Colorado to determine optimal combinations of intervention strategies that would keep the region below a predetermined threshold of SARS-CoV-2 infection density. This analysis used an age-stratified, deterministic SEIR compartmental model of disease transmission, calibrated to cellphone-based mobility data, to simulate infection trajectories during the winter ski season. Under three national infection levels corresponding to high, medium, and low viral importation risk, we estimated the potential impact of interventions including policy and behavior changes, visitor restriction strategies, and case investigation/contact tracing, in order to quantify the relative and absolute impacts of these interventions in the context of the COVID-19 pandemic. Our results suggest that, in the context of low viral importation risk, case investigation/contact tracing and policy and behavior changes may be sufficient to stay below predetermined infection thresholds without visitor restrictions. However, if viral importation risk is high, visitor restrictions and/or screening for infected visitors would be needed to avoid lockdown-like control scenarios and large outbreaks in tourist communities. These findings provide important guidance to tourist destinations for balancing policy impact in future infectious disease outbreaks.

## 1. INTRODUCTION

COVID-19 has profoundly impacted the global landscape through widespread morbidity and mortality, and through damage to economic and social wellbeing. Questions remain on how best to mitigate the fallout resulting from this public health crisis, particularly in the tourism and hospitality sectors, which have suffered from both SARS-CoV-2 itself and the resulting economic restrictions. Small but popular resort destinations have limited public health bandwidth to handle surges in infections and hospitalizations in a pandemic, but also depend on high levels of outside visitation to maintain successful operations and generate essential revenue for the regions where they are located. In Colorado, ski communities struggled not only with high infection rates during the COVID-19 pandemic, but also with economic losses resulting from complete closures during the latter half of the 2019-20 ski season^1^ and restrictions during the 2020-21 ski season. Infectious disease models have been widely used to examine intervention efficacy in the context of the COVID-19 pandemic, but viral transmission in small resort communities, which at times have a majority of their daily population from outside visitors, has not been extensively explored.

These communities face a unique problem for examining both viral transmission and the impacts of control measures. For example, Summit County, Colorado is home to about 31,000 residents year-round,^2^ but this number can more than triple on any given day during the ski season.^3^ With COVID-19, Summit County and other surrounding counties faced two challenges given these unique population dynamics. First, viral importation from outside visitors, beginning with the first case of COVID-19 reported in the state,^4^ raised questions of whether SARS-CoV-2 would be established or re-established in the region throughout the ski season. Second, sudden increases in population density may have resulted in increased crowding and opportunities for viral transmission. While a significant proportion of cases in the Colorado ski regions were among residents and seasonal employees, several outbreaks were linked to adult ski schools, equipment rental shops, restaurants, and lodging accommodations,^5^ all of which were supported significantly by business from outside visitors.

Throughout the pandemic, Colorado ski communities remained popular for locals and tourists alike to escape to the outdoors, with visitation comparable to pre-pandemic levels.^6^ These communities faced many unknowns surrounding transmission dynamics and optimal control measures. One major question for resort communities globally was whether control measures focused on non-residents could make a substantial impact in slowing viral transmission. This study explores the course of SARS-CoV-2 in Colorado’s ski country across a multitude of hypothetical scenarios, and, in an effort to inform future strategies in similar public health crises, estimates the relative impacts of the control measures that were either used or proposed during the 2020-21 ski season.

## 2. METHODS

### 2.1 Study Context

This study focuses on Summit, Eagle, Grand, Pitkin, Routt, and Garfield Counties, a cluster of counties within Colorado that houses multiple major ski resorts and attracts visitors from around the state, country, and world. Table 1 provides the counts of year-round residents in each county by age group based on state demographic data.^7^

**Table 1.** Colorado counties included in this study.

| County | Ski Areas | Resident Population |  |  |  |  |
| --- | --- | --- | --- | --- | --- | --- |
|  |  | Age 0-19 | Age 20-39 | Age 40-64 | Age 65+ | Total |
| Summit | Keystone,<br>Breckenridge,<br>Arapahoe Basin,<br>Copper Mountain | 4,889 | 12,760 | 9,018 | 4,330 | 30,997 |
| Eagle | Vail, Beaver Creek | 12,716 | 17,481 | 18,029 | 6,867 | 55,093 |
| Grand | Winter Park, Ski<br>Granby Ranch | 3,328 | 3,704 | 5,762 | 2,922 | 15,716 |
| Pitkin | Aspen Highlands,<br>Aspen Mountain,<br>Snowmass,<br>Buttermilk | 2,862 | 4,888 | 6,309 | 3,716 | 17,775 |
| Routt | Steamboat,<br>Howelsen Hill Ski<br>Area | 5,718 | 6,981 | 8,811 | 4,128 | 25,638 |
| Garfield | Sunlight Mountain | 15,715 | 16,555 | 19,717 | 8,176 | 60,163 |
| **Total** |  | 45,228 | 62,369 | 67,646 | 30,139 | 205,382 |

This study aims to quantify and compare three interventions for controlling SARS-CoV-2 transmission in this winter destination region: transmission control (TC), case investigation/contact tracing (CI/CT), and visitor restrictions. In this model, TC is used to characterize any combination of behaviors and policies (except CI/CT) that can reduce transmission-relevant contact, or the probability of infection given contact, which can include mask wearing, working from home, school and business closures, capacity limits, maintaining physical distance, socializing outdoors (vs. indoors), and increased hand hygiene practices. The higher the value of TC, the greater the reduction in effective disease transmission, with 0% representing uncontrolled contact rates (akin to the start of the epidemic), and 100% TC representing a complete cessation of transmission-relevant contacts. For reference, a TC level above 90% in this model would be roughly equivalent to the impact of stay-at-home orders.^8^

CI/CT is built into the model by the inclusion of a separate compartment, in which both symptomatic and asymptomatic individuals can be identified and isolated, and potentially exposed individuals can be identified and quarantined. The effectiveness (denoted *τ* in the model) of CI/CT is a product of several components outlined in Table 2. Because public health officials have limited bandwidth to identify and trace cases and their contacts, we compare high levels of CI/CT to no CI/CT in this model under the assumption that after CI/CT capacity is reached, its effectiveness drops dramatically. In reality, the effectiveness of CI/CT drops with increasing caseloads in a more gradual manner than is captured by this model. Visitor restrictions are modeled by halving the number of expected visitors to the region during the ski season as described in Section 2.3.

**Table 2.** Case investigation/contact tracing metrics included in the model. Metrics were estimated by averaging weekly data collected from 33 local public health agencies (LPHA’s) in the state of Colorado between July 13^th^, 2020, and August 10^th^, 2020. Because the reporting LPHA’s varied in size from small to large, estimates of metrics were generated as a weighted average based on the number of weekly cases assigned to each LPHA.

| **Metric** | **Description** | **Value** | **Source** |
| --- | --- | --- | --- |
| $\theta$ | estimated proportion of true infections detected by Colorado state surveillance systems between October 17<sup>th</sup>, 2020 and October 31<sup>st</sup>, 2020 | 0.49 | Colorado COVID-19 Modeling Report, November 12<sup>th</sup>, 2020<sup>9</sup> |
| $\eta$ | average number of contacts successfully contacted per case identified | 0.94 | personal communication, CDPHE |
| $\omega$ | probability that an individual identified through CI/CT is infected | 0.12 | Kucharski, et. al.<sup>10</sup> |
| $\pi$ | probability that an infected individual identified through CI/CT is successfully isolated before infecting others | 0.43 | personal communication, CDPHE |
| $\tau$ | proportion of infectious individuals who isolate and exposed individuals who quarantine as a result of CI/CT in the model | 0.049 | $\eta * \omega * \pi$ |

### 2.2 Transmission Model

SARS-CoV-2 transmission in the region is modeled using a deterministic Susceptible-Exposed-Infected-Recovered (SEIR) model with a dynamic population (Figure 1), stratified into four age groups (0-19, 20-39, 40-64, and 65+) to highlight age-specific differences in COVID-19 severity.^11^ Model equations and parameter values are provided in the Appendix.

**Figure 1.**
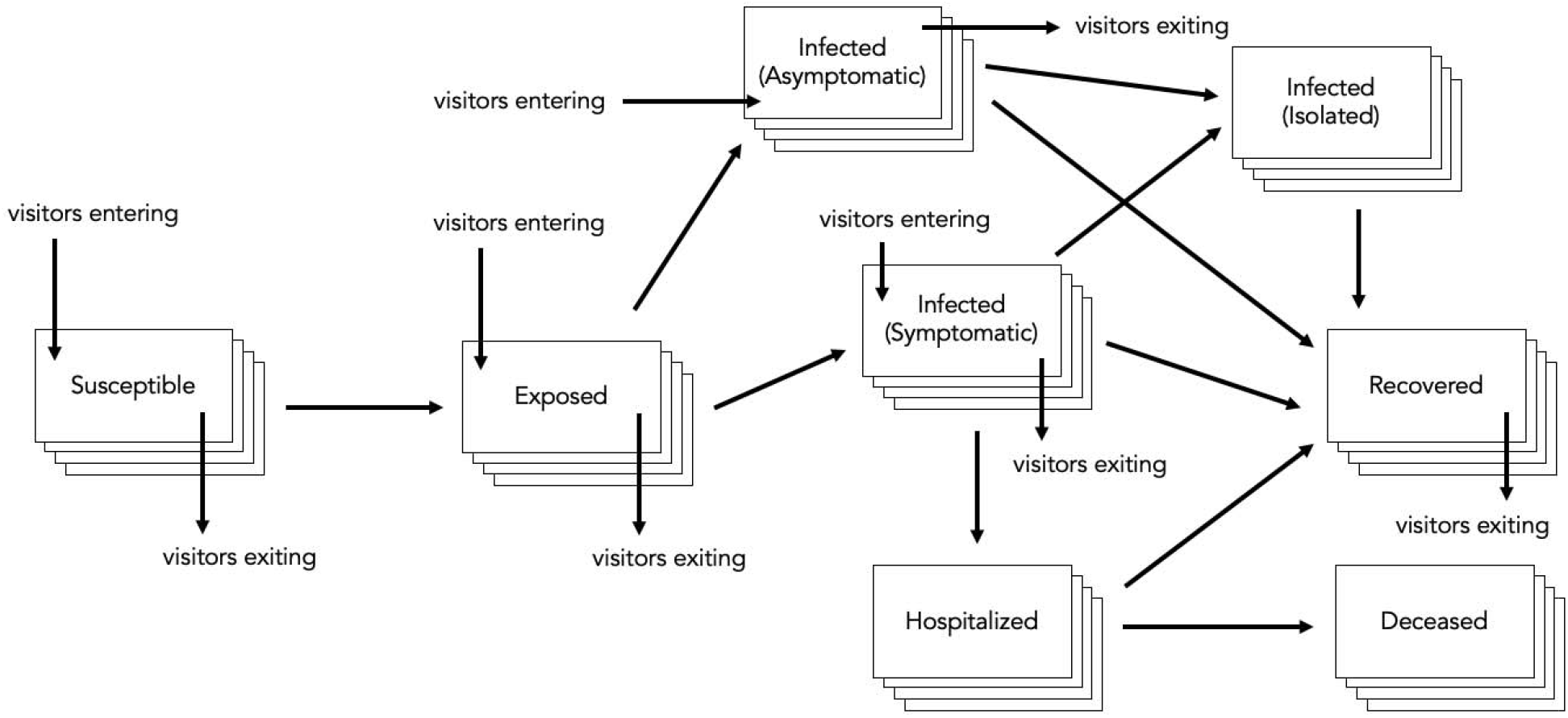
Age-stratified regional SEIR model schematic for this analysis.

### 2.3 Viral Importation Risk

To capture the impact of outside visitation over the course of the ski season, as well as the impact of limiting this outside visitation through visitor restriction, we simulate a dynamic population. Permanent residents of the region are assumed to live in a closed system; we do not consider births, deaths, or travel out of the region among individuals who live there year-round. However, visitors may enter and leave the system freely in this model as indicated by the visitor entry and exit arrows. We estimate visitation from out of the area using data from SafeGraph^12^, a data company that aggregates anonymized location data from numerous applications in order to provide insights about physical places, via the SafeGraph Community.^a^ The data provide estimated counts of visitors to the Colorado ski counties by the devices’ estimated county of origin. SafeGraph reports the number of mobile device visits between all census block groups (CBG) by day in the US. We aggregate daily inter-CBG counts up to the county-level and exclude visits to the county by devices believed to reside in the same county. The result is a daily count of visits from outside the area including Alaska, Hawaii, Washington, D.C., and the rest of Colorado (Figure 2). Visit data from the beginning of the simulation period (January 24^th^, 2020) to the end of the data collection period (November 15^th^, 2020) used the real count of devices given by SafeGraph during this same time frame, which was used when fitting the model throughout the course of the outbreak in Colorado. From November 16^th^, 2020 to April 17^th^, 2021, visit data was abstracted from the previous-year’s SafeGraph data from November 16^th^, 2019 to December 31^st^, 2019, and from January 1^st^, 2019 to April 17^th^, 2019, to mimic visits during an otherwise normal ski season. Under scenarios involving visitor restriction, the count of daily visitors was reduced by 50% beginning December 4^th^, 2020.

**Figure 2.**
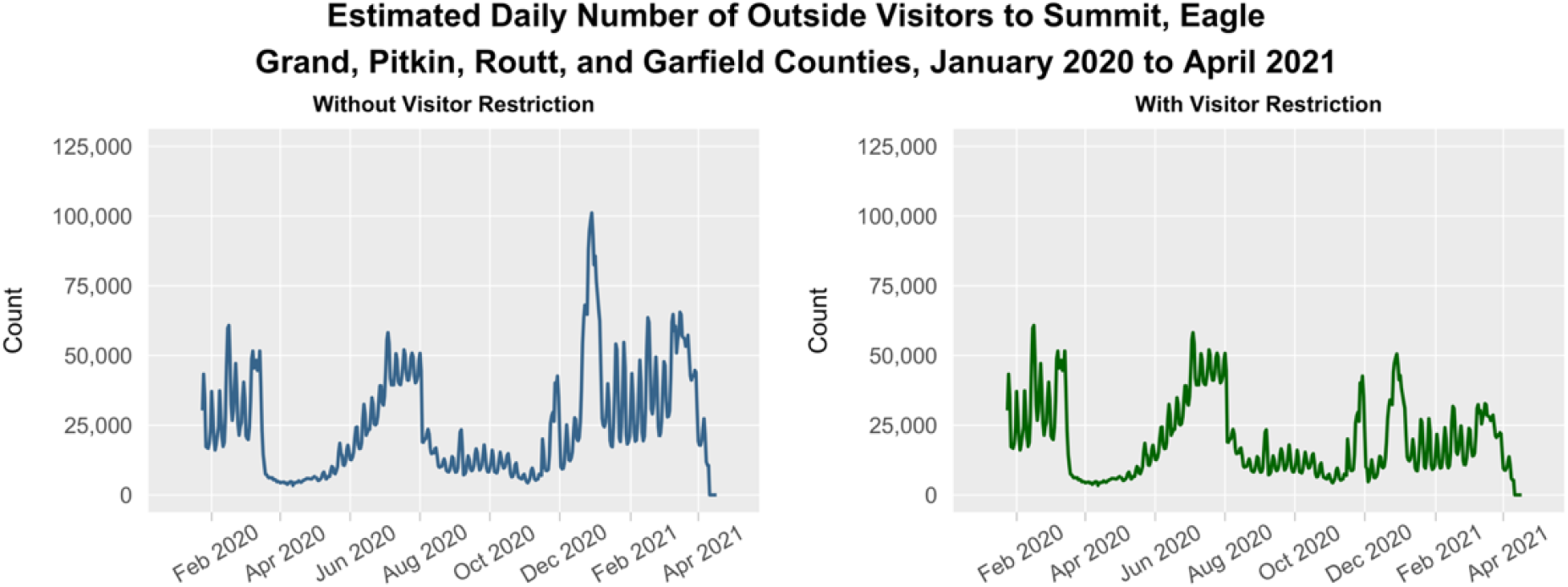
Estimated daily count of outside visitors from January 2020 to April 2021 during a hypothetical normal ski season (left panel). These daily counts are added onto permanent resident population in the model. For scenarios involving a visitor restriction strategy, the daily count of outside visitors for the period spanning December 4^th^, 2020 to April 17^th^, 2021 was reduced by half (right panel).

This count of daily outside visitors is used as a time-varying parameter in the model denoted, assumed to include daytime visitors, vacationing tourists, second homeowners, and seasonal employees in this model. is stratified by age in a manner consistent with the region’s permanent resident age group distribution, where to represent the four age groups, and is added to the existing baseline population at each timestep throughout the simulation. SafeGraph data measures visitor entries, but not the duration of multi-day visits to the region, we assumed that all visitors in the model stayed in the region for an average of one week. Visitors entering the system contribute to the total population and are distributed throughout the Susceptible, Exposed, Infected (Symptomatic), and Infected (Asymptomatic) compartments proportionate to national prevalence estimates under high, medium, and low viral importation risk (Figure 3). A visitor may leave either from the compartment entered or from a downstream compartment, including the Recovered compartment. The Recovered compartment is excluded as a visitor entry point because the impact of visitor immunity through vaccination or natural infection was beyond the scope of this study. All 205,382 permanent residents in the region start the simulation Susceptible, and the model is seeded with one infected individual in the 0-19 age group.

**Figure 3.**
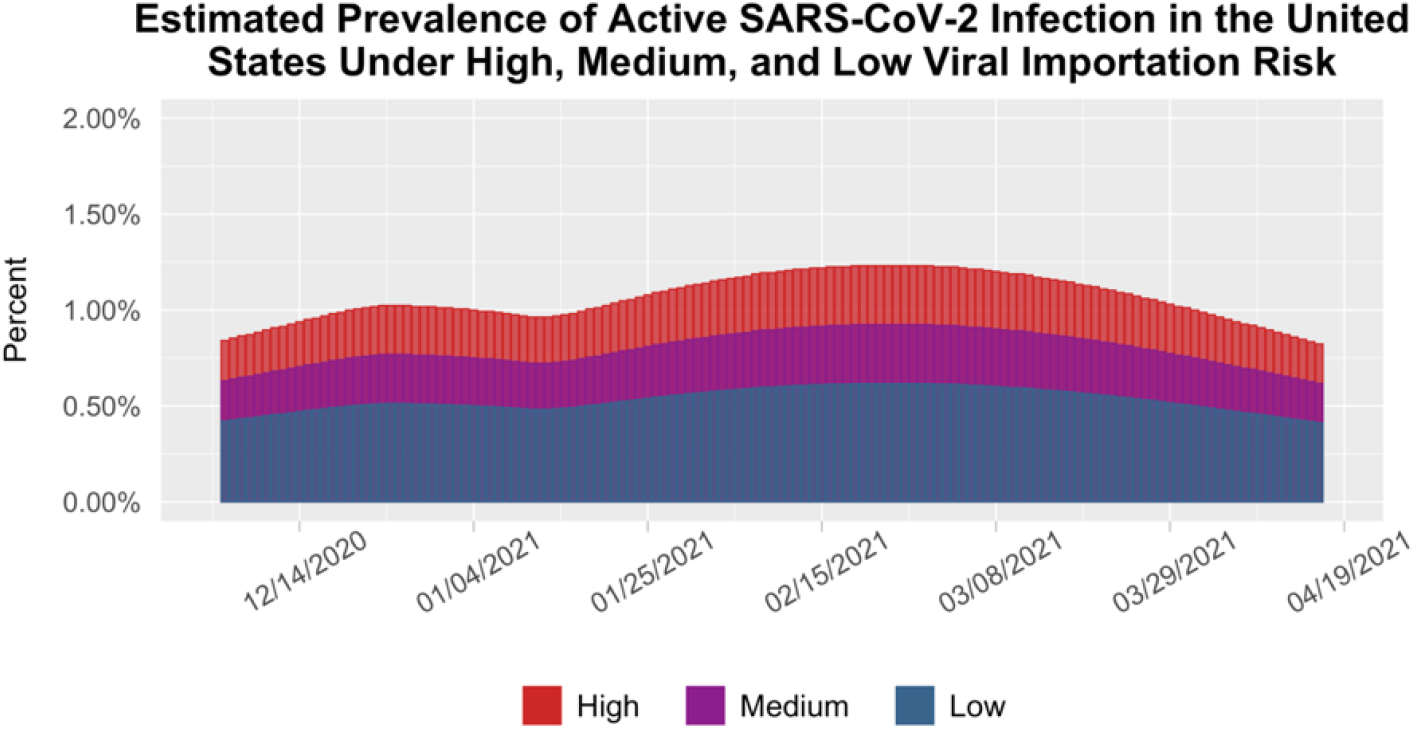
Historical and projected estimates of prevalent SARS-CoV-2 infections in the United States from early December through early May 2021. Estimates are given as a percentage of the total US population.

The variation of TC, visitor restriction, and CI/CT in this study are considered in the context of high, medium, and low viral importation (Figure 3). To ascertain the risk of viral importation into the region for each scenario, denoted in the model, viral importation was derived from historical estimates and future projections of national prevalence of active SARS-CoV-2 infections in the United States from the date of data collection (January 16^th^, 2021) through May 31^st^, 2021 as estimated by the COVID-19 Simulator.^13^ Due to the difference between national prevalence estimates and Colorado prevalence estimates at the time of data collection,^14^ high viral importation scenarios assumed that after December 4^th^, 2020, the percent of visitors infectious (or exposed and soon to be infectious) with SARS-CoV-2 was 2 times higher than national prevalence estimates given by the simulator peaked at 1.2%. Medium viral importation scenarios assumed that after December 4^th^, 2020, the percent of visitors infectious (or exposed and soon to be infectious) with SARS-CoV-2 was 1.5 times higher than national prevalence estimates given by the simulator and peaked at 0.93%. Low viral importation scenarios assumed that the percent of visitors infectious (or exposed and soon to be infectious) with SARS-CoV-2 matched national prevalence estimates given by the simulator, which peaked at 0.62% in mid-February 2021.

### 2.4 Intervention Scenarios

We simulate SARS-CoV-2 transmission in this region during the COVID-19 pandemic in a set of hypothetical scenarios (Table 3), varying the extent of visitor restriction and CI/CT strategies across three levels of viral importation risk from visitors. Viral importation risk is a function of both national prevalence of SARS-CoV-2 infection and the potential use of visitor screening strategies. Under these scenarios, we ascertain the levels of TC needed for infection density in the region to stay below a predetermined threshold. This threshold, known as the Deterministic Incidence Rate Threshold (DIRT), was designed as a benchmark for potentially tolerable levels of transmission. Maintaining transmission low enough to avoid breaching critical care capacity has been a primary public health goal over the course of the pandemic, and in November 2020, working with the Colorado Department of Public Health & Environment (CDPHE), we determined that a two-week model-estimated cumulative incidence density of 2,880 incident SARS-CoV-2 infections (per 100,000) in any given region could translate to a breach of statewide ICU capacity. The DIRT translates into approximately 1,270 reported cases (per 100,000) over this same rolling two-week period, without considering the inherent lag in reporting. In each scenario, we ascertain the minimum level of TC that must be maintained in order for the two-week incidence of new infections in the region to remain below the DIRT for the entire duration of the ski season spanning December 4^th^, 2020 to April 17^th^, 2021.

**Table 3.** Hypothetical combinations of intervention strategies explored under high, medium, and low viral importation scenarios.

| Scenario | Description |
| --- | --- |
| **High Viral Importation** |  |
| 1A | no visitor restriction or CI/CT |
| 1B | CI/CT |
| 2A | visitor restriction |
| 2B | visitor restriction + CI/CT |
| **Medium Viral Importation** |  |
| 3A | no visitor restriction or CI/CT |
| 3B | CI/CT |
| 4A | visitor restriction |
| 4B | visitor restriction + CI/CT |
| **Low Viral Importation** |  |
| 5A | no visitor restriction or CI/CT |
| 5B | CI/CT |
| 6A | visitor restriction |
| 6B | visitor restriction + CI/CT |

### 2.5 Model Fit and Scenario Simulation

This model was fit to daily hospital census data for the six counties obtained from the CDPHE COvid Patient Hospitalization Surveillance (COPHS) database,^15^ which provides a daily estimated count of patients actively hospitalized for COVID-19 in a given ZIP code. To determine the baseline infection rate (*β*) before interventions began, *β* was fit to hospitalizations from January 24^th^ through March 12^th^, 2020, a period characterized by no TC behaviors and followed by an exponential growth phase of the pandemic. After estimating *β*, each time-varying value of *TC* was fit to two-week intervals of data spanning March 13^th^ to December 16^th^, 2020. Estimates of *TC* in this model reflect transmission occurring 13 days prior to the end date of hospitalizations for that period due to the average lag time between initial infection and hospitalization, so the final estimate of *TC* is assumed to reflect transmission occurring on December 3^rd^, 2020. With all other parameters held constant, estimates of each time-varying value of *TC* were obtained using an L-BFGS-B optimization fitting algorithm in R,^16^ giving parameter estimates that yielded the smallest residual sum of squares between the fitted curve and observed number of hospitalizations. Model equations, parameter estimates, and curve fit are provided in the Appendix.

Simulations to determine the minimum levels of TC that would need to be maintained in the region between December 4^th^, 2020, and April 17^th^, 2021, for SARS-CoV-2 infection density to remain below the DIRT for the entire duration of this period, which represents the core ski season, were run under the 12 hypothetical scenarios (Table 3). Model simulations were executed with an ODE solver in R version 1.2.1335.^17^ Simulated TC values after December 4^th^, 2020 for each scenario were chosen from 100 samples of a random uniform distribution ranging from 0.59 to 0.99, for a total of 1,200 runs. The value of 0.59 was used as a lower bound to capture 60% TC, the lowest level seen in the state of Colorado since the end of the stay-at-home order in April 2020 through early 2021.^18^ Incident cases were calculated for each daily timestep using the formula 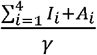. Once all model simulations were complete, the two-week cumulative incidence was calculated by dividing the right-justified 14-day rolling sum of daily incident cases by the baseline population of 205,382 individuals. Each scenario was filtered to yield the lowest possible value of TC that resulted in the two-week cumulative incidence remaining below the DIRT for the entire simulation period.

## 3. RESULTS

Across scenarios, higher levels of outside visitation and viral importation led to an increase in the estimated necessary levels of TC to maintain for the region to remain below the DIRT (Table 4). Notably, visitor restrictions had the greatest impact when viral importation was high. However, when viral importation was low, visitor restrictions had minimal impact, particularly with high levels of CI/CT.

**Table 4.** Minimum TC required for each scenario’s two-week cumulative incidence of SARS-CoV-2 infections (per 100,000 population) to remain below the DIRT for the entire 2020-21 ski season.

| Scenario | Description | $TC_{min}$ |
| --- | --- | --- |
| **High Viral Importation** |  |  |
| 1A | no visitor restriction or CI/CT | 93% |
| 1B | CI/CT | 87% |
| 2A | visitor restriction | 77% |
| 2B | visitor restriction + CI/CT | 71% |
| **Medium Viral Importation** |  |  |
| 3A | no visitor restriction or CI/CT | 82% |
| 3B | CI/CT | 76% |
| 4A | visitor restriction | 73% |
| 4B | visitor restriction + CI/CT | 68% |
| **Low Viral Importation** |  |  |
| 5A | no visitor restriction or CI/CT | 72% |
| 5B | CI/CT | 66% |
| 6A | visitor restriction | 69% |
| 6B | visitor restriction + CI/CT | 63% |

Figure 4 shows the visualization of model-estimated outbreak trajectories for all 12 scenarios. Visitor restriction scenarios actually led to a wider spread of incidence rates across simulations, and sometimes exceeded those in high outside visitation scenarios given small values of *TC*, despite dividing incident cases by the same denominator (the baseline population) in both types of scenarios. This phenomenon occurs in the model because of the homogeneous mixing assumption, which states that interactions in the population system are uniform across space and time. Therefore, a larger number of visitors in the population system actually creates fewer infection moments than a smaller number of visitors if background infection rates are low because although there are more absolute contact moments, each contact moment is less dense, and therefore less transmission-relevant, in the same time span and spatial area in this particular model. In reality, this phenomenon is unlikely to occur in reality because fewer visitors would not necessarily translate directly to denser interactions within this region.

**Figure 4.**
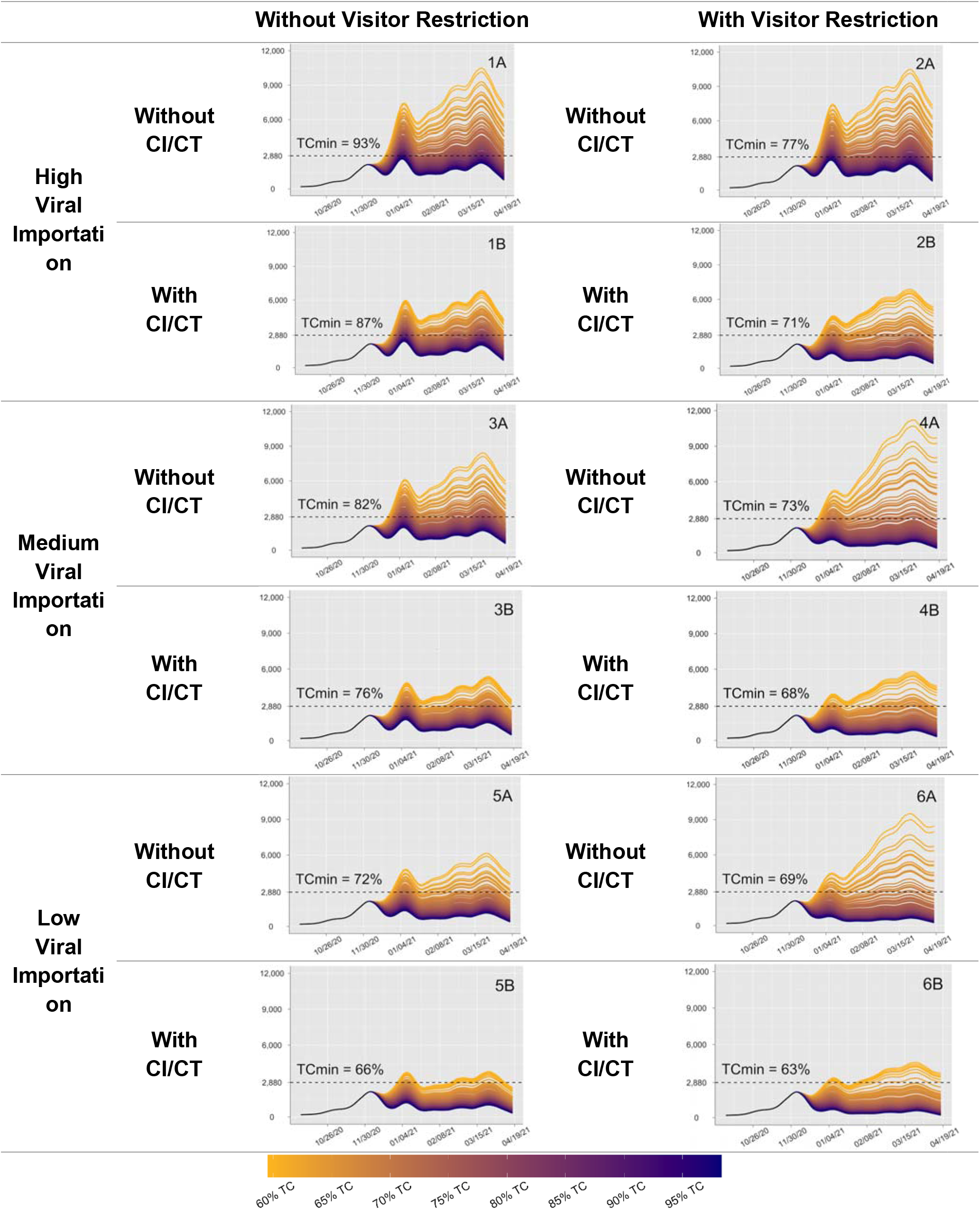
Outbreak trajectories showing model-estimated two-week cumulative incidence (per 100,000 residents using the region’s baseline population as the denominator). Y-axis values represent the right-justified 14-day rolling sum of new infections estimated by the model. The slider scale represents the range of values for from 60% TC to 95% TC. Horizontal dashed lines indicate the DIRT. is provided in the plot area for each scenario.

## 4. DISCUSSION

Using a novel SEIR transmission model with a dynamic population, we estimated the relative value of SARS-CoV-2 control measures for maintaining transmission below a given risk threshold in a tourist region. We found that, in circumstances where national prevalence of SARS-CoV-2 infection is high, and public health officials are unable to implement visitor restrictions or CI/CT, there would need to be near-lockdown conditions (TC ∼93%) in order to keep infection density below the DIRT. This outcome would be undesirable for residents, visitors, public health officials, and business owners alike. On the other side of the spectrum, if viral importation risk is low and both visitor restriction and CI/CT strategies are implemented, TC would need to be maintained only at 63% (a level that would theoretically allow businesses to remain open and function without as much economic strain).

Nevertheless, restriction in outside visitation to the region brings economic strain in and of itself, and policymakers generally consider limiting visitors only when extreme circumstances require. If viral importation risk is low, restricting the number of visitors by half would result in only a marginal relative drop in *TC*_*min*_, meaning that businesses would struggle and infection density would still stay approximately the same. However, if viral importation risk is high, reducing the number of outside visitors by half would result in a substantial relative drop in *TC*_*min*_—essentially, if there is a high chance of visitors arriving infected, then limiting their entry matters. Since restricting visitor entry is an undesirable control measure in a winter destination resort region that is reliant on tourism, more palatable options exist that would allow for lower restriction levels overall. Even if visitor restrictions were not enacted, the region could still potentially reduce viral importation risk through enhanced visitor screening, isolation and quarantine, as well as the use of travel affidavits or proof of a negative COVID-19 test result. The impact of deliberately reducing viral importation risk diminishes if visitor restrictions are already in place. With this in mind, policymakers should consider how visitation during a given season compares to an otherwise normal ski season, especially since aggressive screening policies could discourage even non-infectious visitors from entering the region.

These results are sensitive to a number of limitations and assumptions, some of which are typical of SIR models (i.e., uniform mixing and an inability to characterize the impact of clustered outbreaks or super-spreading events), and others unique to this study. First, the DIRT was estimated as the last level of infection density before a lockdown should be introduced to prevent the state from reaching ICU capacity, a catastrophic outcome. In an ideal situation, infection rates would never approach the DIRT, especially given the high rate of morbidity and mortality that occurs at this level of transmission.

However, the same model framework can be used for different infection thresholds and the results are expected to be consistent regardless of the defined threshold.

Second, across all 12 scenarios, each simulation presumes that the people in the region maintain the same level of transmission control throughout the entire duration of the ski season, regardless of the infection trajectory. In reality, policies and behaviors are likely to evolve as the situation evolves. Known as the “roller coaster” effect, ^19^ people tend to become more vigilant when infections rise and let down their guard when infections fall. While it is possible to parameterize a model with adaptive behavior,^20^ empirically distinguishing voluntary behavior from policy and other factors remains challenging.^21^

Third, the *TC* parameter may be difficult to interpret as it does not discriminate between the different types of additional interventions that can be employed to reduce disease transmission, such as mask wearing, business closures, and social distancing, but rather simply describes the change in transmission compared to the beginning of the epidemic. We chose to collapse all interventions (both policy and behavioral) into a single parameter for two reasons. First, policymakers do not implement control measures individually; they implement many at the same time, often at varying spatial scales. These control measures have an additive effect and are greater than the sum of their parts, making it difficult to accurately estimate the impact of a single intervention. Second, reduction of transmission may look different from one region to the next depending on what leaders and constituents are willing and able to do, so using a single parameter gives some degree of flexibility in how certain strategies may play out. The same notion applies to CI/CT, which consists of several different elements of the contact tracing process that are rolled into a single multiplier representing CI/CT’s ability to blunt uncontrolled transmission. Depending on the public health resources at hand, high levels of CI/CT may mean conducting case interviews more rigorously, identifying more contacts per case, or isolating infectious individuals more quickly.

A fourth limitation was the use of secondary data sources for parameters such as viral importation and CI/CT effectiveness. Estimates from the COVID-19 Simulator, which were used to quantify the risk of viral importation were dependent on the assumptions of their model, which differ in substantive ways from our own model, leading to low estimates of national prevalence overall compared to our own estimates of Colorado prevalence. Another important consideration likely not captured by the COVID-19 Simulator is that skiing and snowboarding are extremely expensive activities, especially for out-of-state visitors who must arrange for travel, lodging, and possible equipment rental or lessons. Visitors that are of higher socioeconomic status have a lower risk of being infectious (or exposed and soon to be infectious) upon entry to the region.^22^ In addition, existing CI/CT data used for this analysis may overestimate its effect in this model, particularly in circumstances of high transmission. CI/CT data were collected over the summer, when SARS-CoV-2 transmission in Colorado was at its lowest levels since the beginning of the pandemic,^23^ and the caseload was relatively manageable. In addition, the data were aggregated for the whole state, and although they were adjusted to account for the proportionate contribution of each county’s reported cases, CI/CT is complicated in the resort communities by high rates of visitors from other states that test positive in-state. Because of their transient nature, a large visitor population inherently decreases the success rate of CI/CT.

Finally, the model does not consider vaccination or the circulation of new, more transmissible variants.^24–25^ Widespread vaccination is expected to allow for substantial reductions in the need for additional control measures. Conversely, because of their increased transmissibility, regular circulation of new variants will drive transmission higher leading to increases in infections, hospitalizations, and deaths without changes to behavior and policy. However, because a majority of the simulation period encompassed a time when vaccines were not yet widely available to the general public, and the spread of variants carried with it a myriad of uncertainties, examining the impact of vaccination and variants was beyond the scope of this study.

In the context of the COVID-19 global pandemic and future global outbreaks, resort communities with high visitation rates may benefit from prioritizing visitor screening and other strategies to reduce viral importation. These strategies should be considered before limiting total visitation levels, as they lead to greater impact at lower economic cost. Preventing viral importation in low population regions may allow tourist destinations to both maintain economic activity with high visitation and circumvent the need for drastic lockdowns and other transmission control measures.

## Data Availability

Model code and all data is available on GitHub: https://github.com/emwu9912/covid-ski

https://github.com/emwu9912/covid-ski

## Funding

This study was supported by the Colorado Department of Public Health and Environment (Contract PO:202000011320-204174) and the Colorado Health Foundation. This content is the sole responsibility of the authors and does not necessarily represent the official views of the funders.

Model code and data is available on GitHub: https://github.com/emwu9912/covid-ski

## Abbreviations

TC: transmission control
CI/CT: case investigation/contact tracing
LPHA: local public health agency
DIRT: deterministic incidence rate threshold
CDPHE: Colorado Department of Public Health & Environment
CBG: census block group
COPHS: COvid Patient Hospitalization Surveillance

## APPENDIX

### SEIR Model Equations and Parameters

The total population of the region, as well as the age-specific populations, are represented by the following sums of all individuals in all compartments of the model at any given time step:

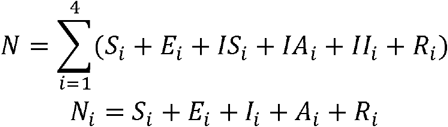

This model is an extension of a previously published model, parameterized to Colorado data and composed of the following system of differential equations, where represents age groups {1, 2, 3, 4}:

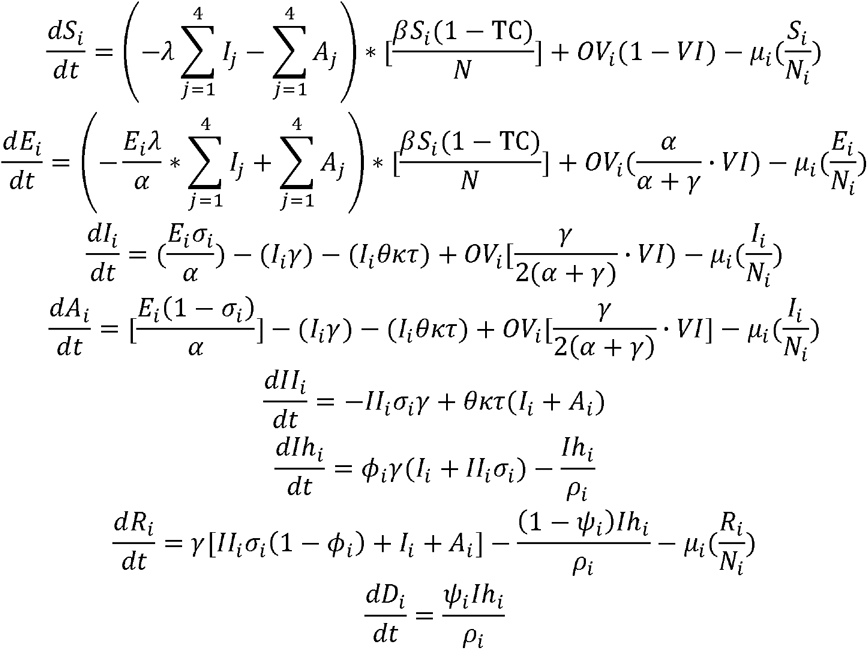

**Table A1.**
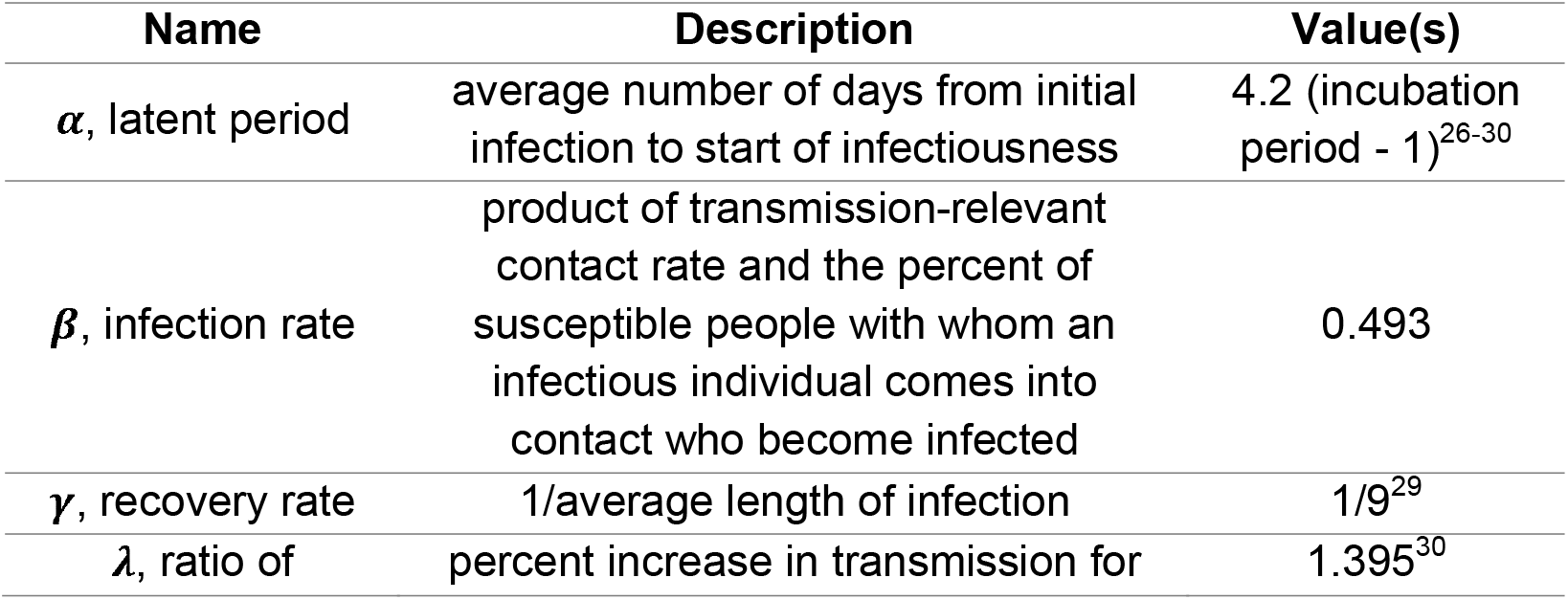
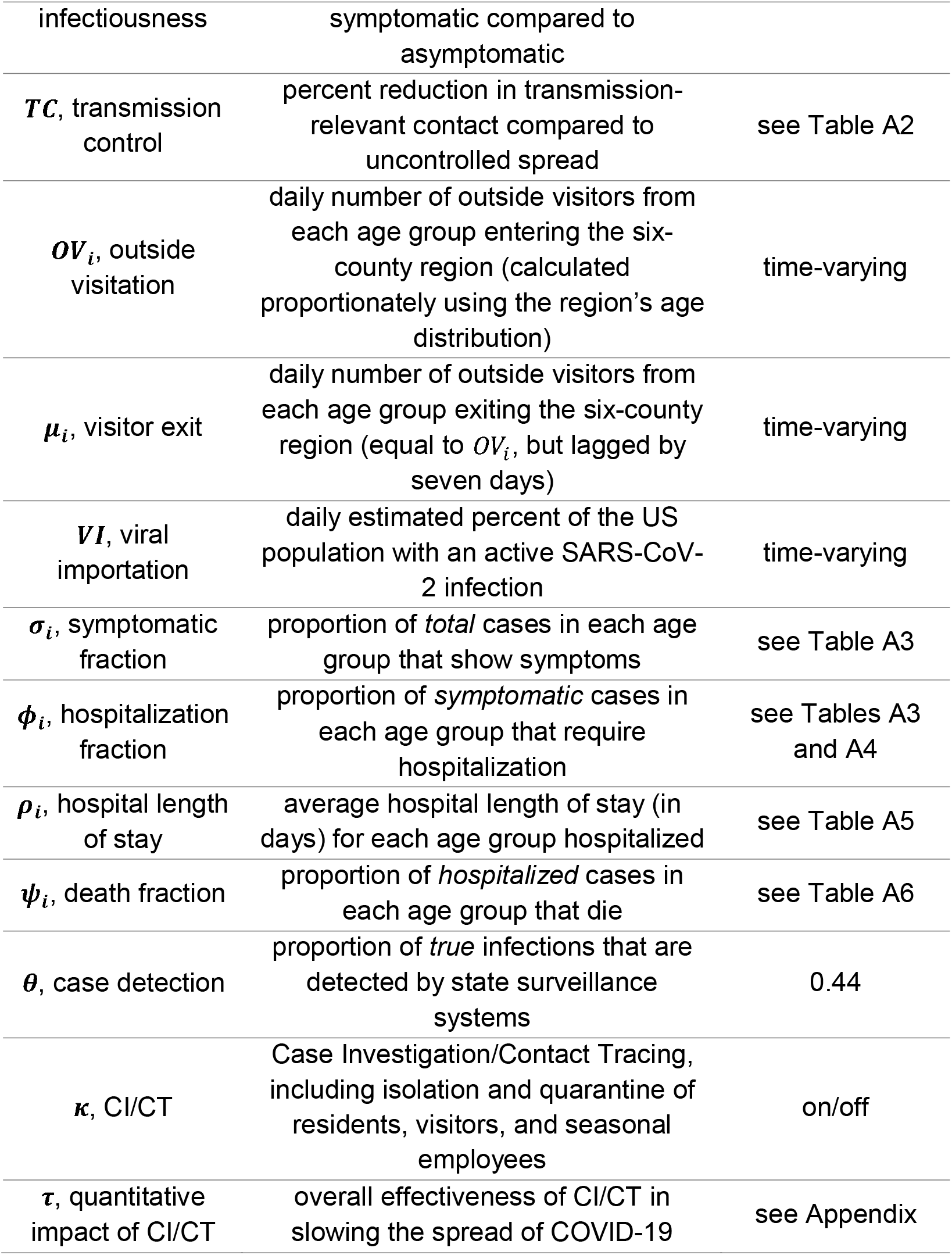
Parameters used in the SEIR model for this analysis.

### Model Fit and Parameter Estimation

**Figure A1.**
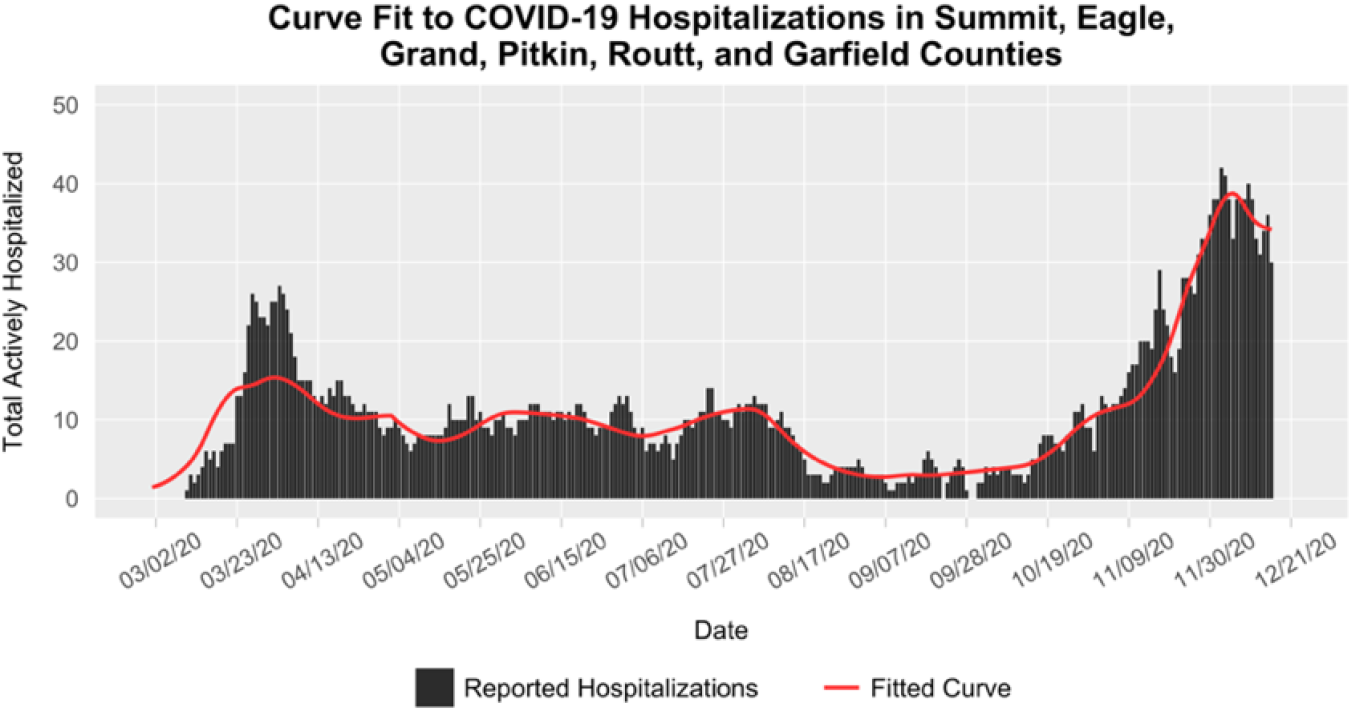
Model estimated hospitalizations fit (red line) to the count of total active COVID-19 hospitalizations (black bars) through December 16^th^, 2020, across the six-county region. Estimates of transmission control were obtained for 20 two-week periods, with the final estimate *TC*_20_ = 0.71 representing transmission control during the two-week period spanning November 20^th^, 2020, to December 3^rd^, 2020.

**Table A2.** Time-varying TC estimates in the six-county region.

| Parameter | Value | TC Date Range | Fit to Hospitalizations Through |
| --- | --- | --- | --- |
|  | 0.0000 | 01/24/2020 – 03/12/2020 | 03/25/2020 |
|  | 0.5868 | 03/13/2020 – 03/26/2020 | 04/08/2020 |
|  | 0.9900 | 03/27/2020 – 04/09/2020 | 04/22/2020 |
|  | 0.7524 | 04/10/2020 – 04/23/2020 | 05/06/2020 |
|  | 0.8496 | 04/24/2020 – 05/07/2020 | 05/20/2020 |
|  | 0.6452 | 05/08/2020 – 05/21/2020 | 06/03/2020 |
|  | 0.7832 | 05/22/2020 – 06/04/2020 | 06/17/2020 |
|  | 0.7717 | 06/05/2020 – 06/18/2020 | 07/01/2020 |
|  | 0.8031 | 06/19/2020 – 07/02/2020 | 07/15/2020 |
|  | 0.6642 | 07/03/2020 – 07/16/2020 | 07/29/2020 |
|  | 0.7959 | 07/17/2020 – 07/30/2020 | 08/12/2020 |
|  | 0.9696 | 07/31/2020 – 08/13/2020 | 08/26/2020 |
|  | 0.9177 | 08/14/2020 – 08/27/2020 | 09/09/2020 |
|  | 0.8150 | 08/28/2020 – 09/10/2020 | 09/23/2020 |
|  | 0.7721 | 09/11/2020 – 09/24/2020 | 10/07/2020 |
|  | 0.7465 | 09/25/2020 – 10/08/2020 | 10/21/2020 |
|  | 0.4958 | 10/09/2020 – 10/22/2020 | 11/04/2020 |
|  | 0.7434 | 10/23/2020 – 11/05/2020 | 11/18/2020 |
|  | 0.5136 | 11/06/2020 – 11/19/2020 | 12/02/2020 |
|  | 0.7146 | 11/20/2020 – 12/03/2020 | 12/16/2020 |
|  | 0.59 –<br>0.999 | 12/04/2020 – 04/17/2021 | projection |

### COVID-19 Clinical Spectrum Data

This model stratifies by age group the proportion of SARS-CoV-2 infections that progress to clinical disease, and the probability of hospitalization and critical care given symptomatic infection (Table A3 and A4).^31^

**Table A3.** Proportion of infections that become symptomatic, and proportion of symptomatic infections that require hospitalization. Estimated from Davies, et. al.^32^

| Age Group | Symptomatic Fraction | Hospitalized Fraction |
| --- | --- | --- |
| 0-19 | 0.11 | 0.0174 |
| 20-39 | 0.36 | 0.0291 |
| 40-64 | 0.56 | 0.0321 |
| 65+ | 0.77 | 0.0501 |

**Table A4.** Proportion of symptomatic infections that require critical care by age group, 2020.

| Age Group | Critical Care Fraction |  |  |
| --- | --- | --- | --- |
|  | Before June 18<sup>th</sup> | June 18<sup>th</sup> to September 13<sup>th</sup> | After September 13<sup>th</sup> |
| 0-19 | 0.00528 | 0.00528 | 0.00528 |
| 20-39 | 0.007474 | 0.00838 | 0.00591 |
| 40-64 | 0.01866 | 0.01785 | 0.012226 |
| 65+ | 0.02885 | 0.02234 | 0.02020 |

In addition, the model considers average ICU and non-ICU hospital length of stay. Mean hospital and ICU lengths of stay were estimated from data collected for 3,947 COVID-19 hospital patients through June 2020, as well as case reports from CDPHE through June 22^nd^, 2020 (Table A5). Mortality was estimated by the Colorado COVID-19 Modeling Group based on data from COVID-19 hospital patients in Colorado and reported COVID-19 case data through September 31^st^, 2020, and using age, hospitalization status, and statewide ICU bed capacity (Table A6).

**Table A5.** Hospital and ICU average length of stay by age group, 2020.

| Age Group | Non-ICU LOS (Days) |  | ICU LOS (Days) |  |
| --- | --- | --- | --- | --- |
|  | Before May 1<sup>st</sup> | After May 1<sup>st</sup> | Before May 1<sup>st</sup> | After May 1<sup>st</sup> |
| 0-19 | 4.05 | 5.0 | 7.0 | 4.0 |
| 20-39 | 5.49 | 3.6 | 9.91 | 4.4 |
| 40-64 | 7.36 | 5.3 | 13.47 | 8.5 |
| 65+ | 10.02 | 7.6 | 10.82 | 8.0 |

**Table A6.** Death fraction for hospitalized (non-ICU) and ICU patients, 2020.

| Age Group | Non-ICU Death Fraction |  | ICU Death Fraction |  |
| --- | --- | --- | --- | --- |
|  | Before July 2<sup>nd</sup> | After July 2<sup>nd</sup> | Before July 2<sup>nd</sup> | After July 2<sup>nd</sup> |
| 0-19 | 0.0000 | 0.0000 | 0.0408 | 0.0408 |
| 20-39 | 0.0000 | 0.0000 | 0.0573 | 0.0573 |
| 40-64 | 0.0100 | 0.0035 | 0.1650 | 0.1101 |
| 65+ | 0.1411 | 0.0609 | 0.3907 | 0.3028 |

<sup>26</sup> Lauer SA, Grantz KH, Bi Q, et al. The Incubation Period of Coronavirus Disease 2019 (COVID-19) From Publicly Reported Confirmed Cases: Estimation and Application. *Annals of Internal Medicine*. 2020;172(9):577-582. doi:10.7326/M20-0504
<sup>27</sup> Tong Z-D, Tang A, Li K-F, et al. Potential Presymptomatic Transmission of SARS-CoV-2, Zhejiang Province, China, 2020. *Emerg Infect Dis*. 2020;26(5):1052-1054. doi:10.3201/eid2605.200198
<sup>28</sup> Linton N, Kobayashi T, Yang Y, et al. Incubation Period and Other Epidemiological Characteristics of 2019 Novel Coronavirus Infections with Right Truncation: A Statistical Analysis of Publicly Available Case Data. *JCM*. 2020;9(2):538. doi:10.3390/jcm9020538
<sup>29</sup> *Modeling Update: The Current State of COVID-19 in Colorado*. Colorado COVID-19 Modeling Group; February 3, 2021.
<sup>30</sup> Zou L, Ruan F, Huang M, et al. SARS-CoV-2 Viral Load in Upper Respiratory Specimens of Infected Patients. *N Engl J Med*. 2020;382(12):1177-1179. doi:10.1056/NEJMc2001737

## Footnotes

a To enhance privacy, SafeGraph excludes census block group information if fewer than two devices visited an establishment in a month from a given census block group.

